# Estimation of case-fatality rate in COVID-19 patients with hypertension and diabetes mellitus in the New York State

**DOI:** 10.1101/2020.08.20.20178962

**Authors:** Yang Ge, Shengzhi Sun, Ye Shen

## Abstract

We estimated the case-fatality rate (CFR) and ratios (RR) in adult COVID-19 cases with hypertension and diabetes mellitus in the New York State. We found that the elderly population had a higher CFR, but the elevated CFR ratios associated with comorbidities are more pronounced for the younger population.

---

As of May 18, coronavirus disease 2019 (COVID-19) has affected more than 200 countries and regions^1^. The New York state was heavily impacted by the coronavirus with more than 22 thousands of deaths^2^. Prior studies suggest that pre-existing health conditions such as hypertension (HBP) and diabetes mellitus (DM) may exacerbate the severity of COVID-19^3^. Quantitative assessments of the risk of death associated with them can inform the disease control globally. However, few studies have estimated the corresponding case fatality rate for these two comorbidities.

In this study, we aim to calculate the case-fatality rate (CFR) in adult COVID-19 cases with hypertension and diabetes mellitus based on the publicly reported COVID-19 tracker data in the New York State.

State-wide individual-level data of COVID-19 cases are not publicly available currently. In this study, we aim to estimate CFR in COVID-19 patients with hypertension and diabetes mellitus using aggregated level data from multiple sources. We first obtained the total number of COVID-19 cases and deaths from public reports provided by the New York State Department of Health^2^. For each comorbidity, we then used the corresponding age-specific prevalence reported from the Behavioral Risk Factor Surveillance System 2017 (BRFSS)^4^.

We defined the CFR of COVID-19 cases with one comorbidity as the total number of deaths with the comorbidity divided by the total number of COVID-19 confirmed cases with that comorbidity. To further control for age, we performed stratification and calculated the CFR by different age groups (18–44 y, 45–64 y, and 65+ y).

We obtained the total number of COVID-19 deaths, deaths with comorbidities of hypertension (D_HBP_) or diabetes mellitus (D_DM_), and the total number of confirmed cases from *the New York State Department of Health COVID-19 Tracker*^2^. The tracker, updated daily, includes counts of confirmed cases from all lab testing samples, and age group-specific counts of deaths related to comorbidities reported by health care facilities.

The counts of confirmed cases were reported without age group information in New York State, so we estimated the age group-specific number of cases according to the age distribution of confirmed COVID-19 cases in the contiguous US reported from the *Centers for Disease Control and Prevention*^5^. In addition, information on comorbidities at an aggregate level was only reported for deaths but not all confirmed cases from the Tracker. To estimate the number of cases with hypertension (C_HBP_) or diabetes mellitus (C_DM_), we multiplied the counts of confirmed cases by the corresponding prevalence of hypertension and diabetes mellitus in the New York State from BRFSS^4^ 2017, assuming that the susceptibility of COVID-19 infection for patients of these two comorbidities is similar to the general population. More details are provided in the supplementary material.

Subsequently, we calculated the CFR for hypertension (CFR_HBP_) or diabetes (CFR_DM_) in each age group. We further obtained the CFR ratio (RR) to compare the risk of COVID-19 death with and without the comorbidities.

We restricted our analyses to COVID-19 deaths aged 18 years and above, which included 340,338 (96.9% of all cases) confirmed cases and 22,708 (99.9% of all deaths) deaths by the date of May 18 in the New York State. Hypertension and diabetes mellitus in the New York State were two common comorbidities of COVID-19 cases with the estimated prevalence of 32.2% (95% CI: 30.4%, 34.0%) and 11.9% (95% CI: 10.6%, 13.2%), respectively. As expected, the prevalence of hypertension (Figure 1A) and diabetes mellitus (Figure 1B) in COVID-19 cases was highest among patients aged 65 years and above, with the case-fatality rate being 21.08% (95% CI: 20.25%, 21.99%) for hypertension (Figure 1C) and 30.53% (95% CI: 28.11%, 33.40%) for diabetes mellitus (Figure 1D). Compared with COVID-19 patients who were free of hypertension, those with hypertension are associated with higher case-fatality rate ratios at 2.6 (95% CI: 2.3, 2.9), 1.7 (95% CI: 1.5, 1.8), and 1.0 (95% CI: 0.9, 1.1) for patients aged 18–44, 45–64, and 65+ y, respectively. The corresponding case-fatality rate ratios for diabetes mellitus were 15.2 (95% CI: 12.2, 19.9), 4.1 (95% CI: 3.6, 4.6), and 1.7 (95%CI: 1.5, 1.9), respectively (Figure 1E).

**Figure 1:**
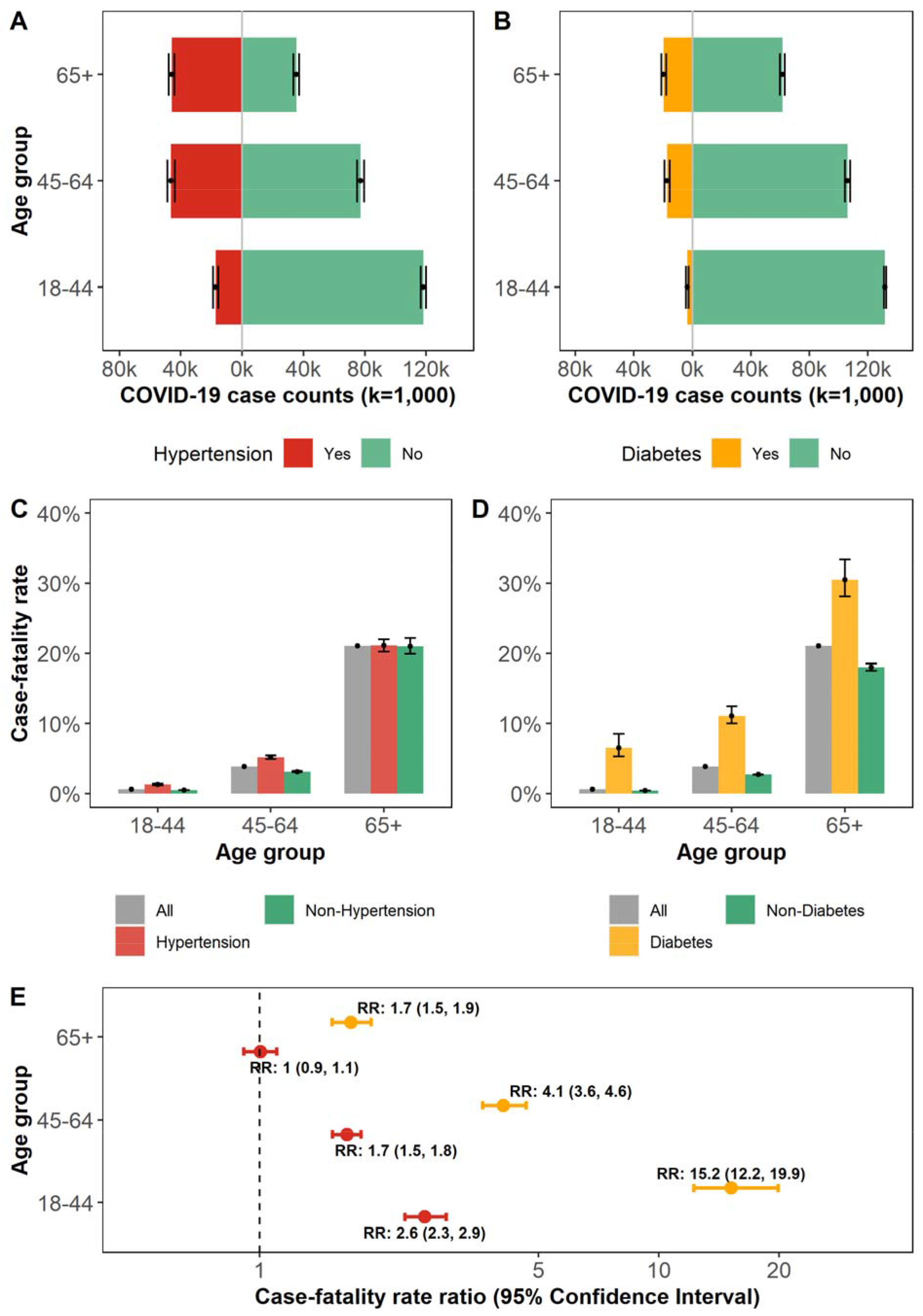
Estimation of the case-fatality rate in COVID-19 patients with hypertension or diabetes. **A**: Age distribution of confirmed COVID-19 cases with hypertension. **B**: Age distribution of confirmed COVID-19 cases with diabetes mellitus. **C**: Age group-specific case-fatality rate among COVID-19 patients in all cases, with or without hypertension, respectively. **D**: The COVID-19 age group-specific case-fatality rate in all cases, with or without diabetes, respectively. **E:** The age group-specific case-fatality rate ratios (RR) of COVID-19 cases with hypertension or diabetes.

We found that COVID-19 patients with hypertension and diabetes were associated with an increased risk of death in New York State. While the elderly population had a higher CFR, the elevated CFR ratios associated with comorbidities are more pronounced for the younger population.

Our results should be considered as preliminary estimations with several limitations. The data provided by the New York State Department of Health COVID-19 Tracker was at an aggregated level with inconsistent stratifications of age. To calculate age group-specific results, we relied on information from data sources of CDC and BRFSS to estimate age distribution and comorbidity prevalence. Those estimations may be biased. In addition, with no individual-level data available for the analyses, further investigations such as the interactions between comorbidities were not conducted.

## Data Availability

https://coronavirus.health.ny.gov

## About the Author

Yang Ge is a Ph.D. candidate of Epidemiology and Biostatistics at the University of Georgia, Athens, Georgia, USA. His areas of interest and research are infectious disease modeling and optimization of vaccines.

## Address for correspondence

Yang Ge, Department of Epidemiology and Biostatistics, University of Georgia, 101 Buck Rd., Athens 30602–7396, Georgia, USA.

Shen Ye, Department of Epidemiology and Biostatistics, University of Georgia, 101 Buck Rd., Athens 30602–7396, Georgia, USA.

## Supplementary Material

**Table S1.** Key data for case-fatality calculation

| Age Group | From the New York State Department of Health |  |  |  | From Behavioral Risk Factor Surveillance System (2017) *2 |  |
| --- | --- | --- | --- | --- | --- | --- |
|  | Confirmed cases*1 | Total deaths | Death with hypertension (D<sub>HBP</sub>) | Death with diabetes (D<sub>DM</sub>) | Hypertension prevalence (%) | Diabetes prevalence (%) |
| 18-44 | 135430 | 795 | 218 | 228 | 12.70 (11.38, 14.03) | 2.58 (1.98, 3.18) |
| 45-64 | 123600 | 4815 | 2399 | 1926 | 37.51 (35.58, 39.43) | 14.04 (12.54, 15.54) |
| 65+ | 81308 | 17098 | 9688 | 5972 | 56.52 (54.19, 58.85) | 24.06 (21.99, 26.13) |
| Total | 340338 | 22708 | 12305 | 8126 |  |  |

\*1: The age distribution of confirmed cases was estimated from the age distribution of confirmed cases in the contiguous US reported by the Centers for Disease Control and Prevention<sup>5</sup>.
\*2: In the calculation of prevalence, pregnancy-related hypertension or diabetes mellitus were excluded.
COVID-19 cases with hypertension (C<sub>HBP</sub>) = Confirmed Cases × Hypertension prevalence
COVID-19 cases with diabetes (C<sub>DM</sub>) = Confirmed Cases × Diabetes prevalence
For each age group, we calculated the case-fatality rate and rate ratios with the following formulas:
CFR<sub>HBP</sub>: case-fatality rate of COVID-19 with hypertension
CFR<sub>Non-HBP</sub>: case-fatality rate of COVID-19 without hypertension
RR<sub>HBP</sub>: case-fatality rate ratio of COVID-19 with hypertension
CFR<sub>DM</sub>: COVID-19 case-fatality rate of COVID-19 with diabetes
CFR<sub>Non-DM</sub>: case-fatality rate of COVID-19 without diabetes
RR<sub>DM</sub>: case-fatality rate ratio of COVID-19 with diabetes

